# Multi-Organ System Metabolic Stress and Sex-Divergent Vascular Associations

**DOI:** 10.1101/2024.05.06.24306949

**Authors:** Alan C. Kwan, Minhao Wang, Hongwei Ji, Brian Claggett, David Ouyang, Hirsh Trivedi, Sonia Sharma, John Shyy, Amanda Velazquez, Joseph E. Ebinger, Susan Cheng

## Abstract

**Introduction:** Women experience excess cardiovascular risk compared to men in the setting of similar metabolic disease burden. This consistent finding could be related to sex differences in the vascular response to various forms of metabolic stress. In this study we examine the association of both systemic and organ-specific metabolic stress with vascular health in women and men.

**Methods:** We conducted an observational study of 4,299 adult participants (52% women, aged 59±13 years) of the National Health and Nutrition Examination Survey (NHANES) 2017-2018 cohort and 110,225 adult outpatients (55% women, aged 64±16 years) of the Cedars-Sinai Medical Center (CSMC) 2019 cohort. We used natural splines to examine the association of systemic and organ-specific measures of metabolic stress including body mass index (BMI), hemoglobin A1c (HbA1c), hepatic FIB-4 score, and CKD-EPI estimated glomerular filtration rate (eGFR) on systolic blood pressure (SBP). Piecewise linear models were generated using normal value thresholds (BMI <25 kg/m^2^, HbA1c <5.7%, FIB-4 <1.3, and eGFR ≥90 ml/min), which approximated observed spline breakpoints. The primary outcome was increase in SBP (relative to a sex-specific physiologic baseline SBP) in association with increase in level of each metabolic measure.

**Results:** Women compared to men demonstrated larger magnitudes and an earlier onset of increase in SBP per increment increase across all metabolic stress measures. The slope of SBP increase per increment of each metabolic measure was greater for women than men particularly for metabolic measures within the normal range, with slope differences of 1.71 mmHg per kg/m2 of BMI, 9.61 mmHg per %HbA1c, 6.45 mmHg per FIB-4 unit, and 0.37 mmHg per ml/min decrement of eGFR in the NHANES cohort (P difference <0.05 for all). Overall results were consistent in the CSMC cohort.

**Conclusions:** Women exhibited greater vascular sensitivity in the setting of multiple types of metabolic stress, particularly in periods representing the transition from metabolic health to disease. These findings underscore the importance of involving early metabolic health interventions as part of efforts to mitigate vascular risks in both women and men.

## Introduction

Decades of clinical trial and observational study data have consistently shown that women experience greater cardiovascular disease (CVD) risk than men in the setting of obesity, insulin resistance, and Type 2 diabetes.^1,2^ The reasons for this persistent sex disparity remain unclear. While theoretically attributable to more frequent clustering of metabolic traits in women than men, the excess risk in women is seen to persist even after adjustment for these factors.^3,4^ Intriguingly, recent studies have suggested the vascular response to similar doses of chronic metabolic stress may be more pronounced in females than males over the life course.^5,6^ Given that aggregate metabolic stress may arise from disease involving one or more distinct organ systems – including the liver and kidney – it is possible that associated excess vascular risk is not only sex biased but also predominantly related to a particular metabolic organ system or systemic process. To investigate this possibility, which would inform treatment approaches, we examined sex-specific systolic blood pressure (SBP) relations with graded severity of organ-specific and systemic measures of metabolic stress.

## Methods

We conducted our primary analyses using adult participant data from the National Health and Nutrition Examination Survey (NHANES) 2017-2018 cohort, representing the most recent complete pre-pandemic NHANES dataset available. To focus on adults at risk of subclinical CVD, we limited the population to adults age ≥35 years, with systolic blood pressure (SBP) between 100 mmHg and 180 mmHg. We selected widely accessible systemic and organ-based measures of metabolic stress including: body mass index (BMI), hemoglobin A1c (HbA1c), FIB-4 (a marker of subclinical hepatic disease commonly assessed in the setting of steatotic liver disease),^7^ and estimated glomerular filtration rate (eGFR) calculated using the 2021 CKD-EPI formula.^8^ We used natural splines to examine SBP in relation to each metabolic stress measure, with 4 knots and 95% confidence intervals, excluding outliers beyond the <2.5% and >97.5% range to avoid extreme deviations in the spline endpoints. We then used piecewise linear splines with a single knot to examine the relations above and below normal value thresholds (BMI of 25 kg/m^2^, HbA1c of 5.7%, FIB-4 of 1.3, and GFR of 90 ml/min), comparing the slopes between women and men for each linear segment using interaction terms. Measures of SBP increase are presented in the splines as the difference in SBP (i.e. SBP shift) from the sex-specific normal healthy (i.e. physiologic) thresholds (110 mmHg for women, 120 mmHg for men) previously established from multi-cohort epidemiologic reference data.^9^

For external validation cohort analyses, we curated outpatient data from Cedars-Sinai Medical Center (CSMC), a large urban academic medical center, between January 1, 2019 and December 31, 2019; we included data from all ambulatory patients with a recorded outpatient blood pressure and laboratory draw involving a complete blood count and comprehensive metabolic panel. If available, HbA1c measures were collected. If multiple measurements were recorded, the average value (for SBP or laboratory measure) was considered representative for that year. Demographic data including race and ethnicity were self-reported during visits, and comorbidities were obtained from the electronic health record via *ICD-10* codes (**Supplemental Table 1**). Ethical approval was obtained for both cohorts (**Supplement**); analyses were conducted using R v4.3.1, and STATA-SE 14; a two-tailed P<0.05 was considered significant.

## Results

Our NHANES community-based sample included 4,299 unique adult participants (52% women, mean age 59±13 years). Self-reported comorbidities included 46% with history of hypertension, 43% with hyperlipidemia, 20% with diabetes, and 6% with coronary artery disease. The CSMC patient-based sample included 110,225 unique adult patients (55% women, mean age 64±16 years) (**Table 1**). Documented comorbidities included hypertension in 40% of patients, hyperlipidemia in 46%, diabetes in 14%, and coronary artery disease in 2.7%.

**Table 1.** Cohort Characteristics. NHANES: National Health and Nutrition Examination Survey; CSMC: Cedars-Sinai Medical Center; HbA1c: Hemoglobin A1c; eGFR: Estimated glomerular filtration rate.

| Characteristics | Community-Based<br>NHANES Cohort Sample |  |  | Ambulatory Patient-Based<br>CSMC Cohort Sample |  |  |
| --- | --- | --- | --- | --- | --- | --- |
|  | Women<br>N=2,214 | Men<br>N=2,085 | P value | Women<br>N=60,823 | Men<br>N=48,402 | P value |
| Age, years | 58±14 | 59±13 | 0.017 | 64±16 | 64±16 | 0.075 |
| Race/Ethnicity |  |  | 0.032 |  |  | <0.001 |
| White | 765 (35%) | 785 (38%) |  | 35,812 (59%) | 31,659 (65%) |  |
| Black | 525 (24%) | 484 (23%) |  | 7,252 (12%) | 3,924 (8.1%) |  |
| Asian | 330 (15%) | 276 (13%) |  | 6,185 (10%) | 3,763 (7.8%) |  |
| Hispanic | 508 (23%) | 431 (21%) |  | 6,884 (11%) | 4,657 (9.6%) |  |
| Other | 86 (3.9%) | 109 (5.2%) |  | 4,690 (7.7%) | 4,399 (9.1%) |  |
| Systolic blood pressure, mmHg | 131±22 | 131±19 | 0.5 | 125±14 | 127±13 | <0.001 |
| Diastolic blood pressure, mmHg | 72±13 | 75±13 | <0.001 | 74±9 | 76±9 | <0.001 |
| Body mass index, kg/m<sup>2</sup> | 31±8 | 29±6 | 0.002 | 26.8±6.5 | 27.7±5.1 | <0.001 |
| HbA1c, % | 5.98±1.13 | 6.08±1.20 | <0.001 | 5.79±0.96 | 5.94±1.11 | <0.001 |
| FIB-4 | 1.24±0.75 | 1.45±1.21 | <0.001 | 1.33±1.01 | 1.53±1.24 | <0.001 |
| eGFR, ml/min | 89±22 | 86±21 | <0.001 | 83±22 | 80±22 | <0.001 |
| Comorbidities, % |  |  |  |  |  |  |
| Hypertension | 1,013 (46%) | 971 (47%) | 0.3 | 22,158 (36%) | 21,670 (45%) | <0.001 |
| Hyperlipidemia | 931 (42%) | 910 (44%) | 0.3 | 24,817 (41%) | 25,896 (54%) | <0.001 |
| Diabetes | 389 (18%) | 469 (22%) | <0.001 | 7,346 (12%) | 8,155 (17%) | <0.001 |
| Coronary Artery Disease | 72 (3.3%) | 192 (9.2%) | <0.001 | 1,085 (1.8%) | 1,896 (3.9%) | <0.001 |

Overall, women exhibited a greater magnitude of increased SBP in association with increase in each measure of metabolic stress (**Figure**). Expectedly, the rate of SBP increase was highest in a period of presumed transition from metabolic health to metabolic disease, prior to when any given measure exceeded the normal range. Notably, the slope of SBP increase in this pre-clinical period was significantly higher for women than men across all metabolic stress measures in the primary NHANES cohort (**Figure** and **Table 2**), with prominent sex differences seen for BMI (β [SE] was 0.99 [0.34] in women, -0.72 [0.33] in men), HbA1c (15.79 [2.07] in women, 6.18 [1.83] in men), FIB-4 (17.76 [3.25] in women, 11.35 [1.74] in men), and eGFR (0.49 [0.06] in women, 0.12 [0.06] in men) (P sex difference <0.05 for all). In the ranges consistent with overt metabolic disease, elevations in SBP with increasing measures of metabolic stress were generally less steep with attenuated sex differences, although the sex differences remained significant for FIB-4 (**Figure** and **Table 2**). In the CSMC patient cohort, the same analyses revealed steeper SBP slopes for women than men across both normal and abnormal ranges of HbA1c, FIB-4, and eGFR; the only exception to this trend was the observation of parallel SBP slopes (i.e. no sex difference) seen for both the normal and abnormal ranges of BMI.

**Figure.**
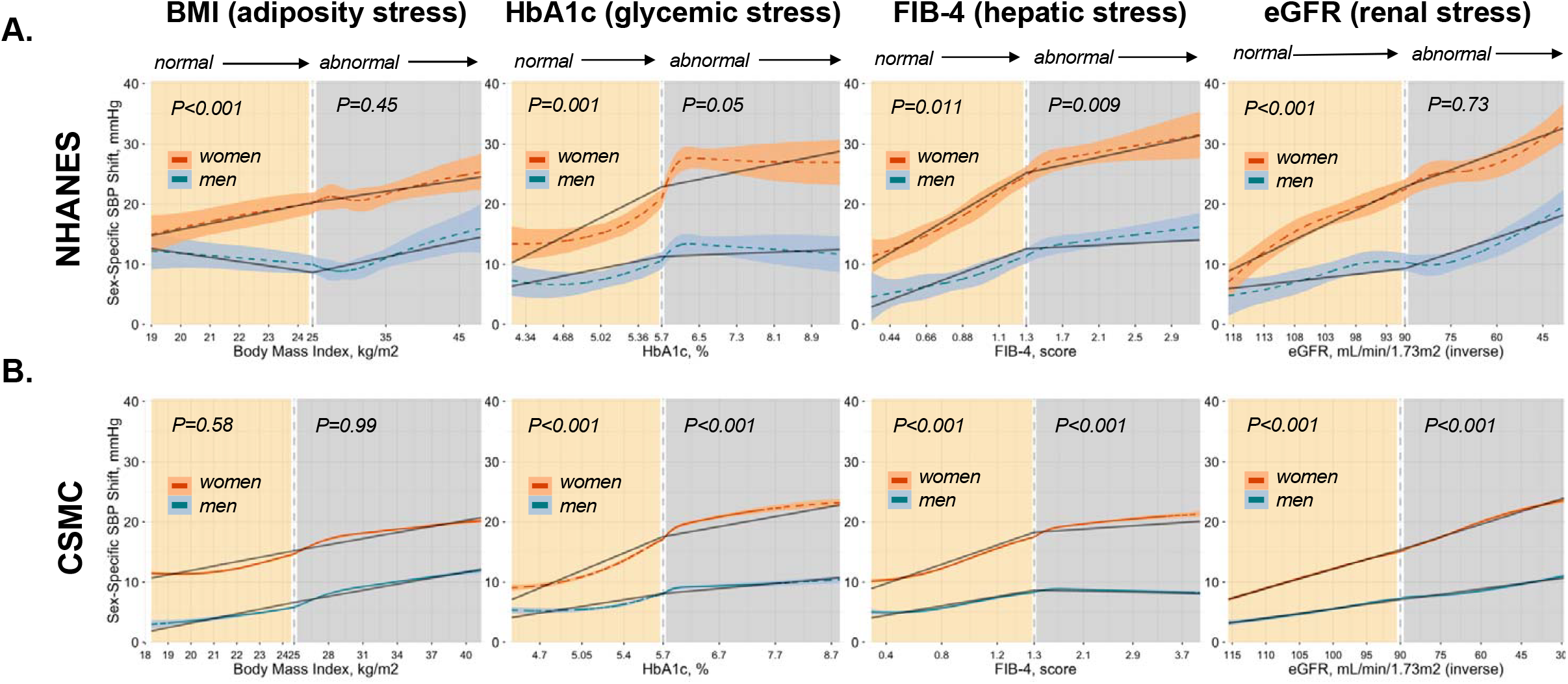
Multi-organ system metabolic stress measures in relation to vascular health, by sex. P values are shown for the sex difference in the segmented slope of rise in systolic blood pressure (SBP), serving as a measure of vascular health, across each measure of metabolic stress categorized as within the normal or with the abnormal range for each measure. Women demonstrate both earlier-onset and greater vascular sensitivity to accumulation of metabolic stress of various types, as reflected by a significantly steeper initial rise in SBP from sex-specific normal healthy (i.e. physiologic) thresholds (110 mmHg for women, 120 mmHg for men) previously established from multi-cohort epidemiologic reference data. NHANES: National Health and Nutrition Examination Survey; CSMC: Cedars-Sinai Medical Center; BMI: Body mass index; HbA1c: Hemoglobin A1c; eGFR: Estimated glomerular filtration rate.

**Table 2.** Associations of metabolic stress measures with systolic blood pressure, by sex. Linear piecewise regression coefficients represent the slope of association between a given metabolic stress measure and increase in systolic blood pressure (SBP) above and below the normal value thresholds, by sex. P values are shown for difference in regression slope between women and men, within the normal or abnormal range of values for each metabolic measure, as calculated in the NHANES community-based cohort and CSMC patient-based cohort samples. Women exhibit steeper slope associations of metabolic stress measures with SBP than men, particularly when assessed within the normal range of values for a given metabolic measures. Bold face denotes P<0.05 levels of statistical significance. NHANES: National Health and Nutrition Examination Survey; CSMC: Cedars-Sinai Medical Center; BMI: Body mass index; HbA1c: Hemoglobin A1c; eGFR: Estimated glomerular filtration rate.

| Metabolic stress measure (normal value threshold) | Community-Based<br>NHANES Cohort Sample (N=4,299) |  |  |  |  |  | Ambulatory Patient-Based<br>CSMC Cohort Sample (N=109,225) |  |  |  |  |  |
| --- | --- | --- | --- | --- | --- | --- | --- | --- | --- | --- | --- | --- |
|  | Within Sex Piecewise Linear Regression Coefficients |  |  |  | Between Sex Comparisons |  | Within Sex Piecewise Linear Regression Coefficients |  |  |  | Between Sex Comparisons |  |
|  | Women (N=2,214) |  | Men (N=2,085) |  | Normal Range | Abnormal Range | Women (N=60,823) |  | Men (N=48,402) |  | Normal Range | Abnormal Range |
|  | Normal Range<br>Slope est.<br>(SE) | Abnormal Range<br>Slope est.<br>(SE) | Normal Range<br>Slope est.<br>(SE) | Abnormal Range<br>Slope est.<br>(SE) | P diff in<br>slope in<br>women vs<br>men | P diff in<br>slope in<br>women<br>vs men | Normal Range<br>Slope est.<br>(SE) | Abnormal Range<br>Slope est.<br>(SE) | Normal Range<br>Slope est.<br>(SE) | Abnormal Range<br>Slope est.<br>(SE) | P diff in<br>slope in<br>women<br>vs men | P diff in<br>slope in<br>women<br>vs men |
| BMI (<25 vs ≥25) | **0.99 (0.34)** | **0.18 (0.07)** | **-0.72 (0.33)** | **0.26 (0.09)** | **<0.001** | 0.538 | **0.73 (0.03)** | **0.34 (0.01)** | **0.76 (0.05)** | **0.34 (0.01)** | 0.594 | 0.988 |
| HbA1c (<5.7 vs ≥5.7) | **15.79 (2.07)** | **1.55 (0.50)** | **6.18 (1.83)** | 0.30 (0.42) | **0.001** | 0.053 | **11.62 (0.31)** | **1.67 (0.10)** | **4.40 (0.33)** | **0.84 (0.09)** | **<0.001** | **<0.001** |
| Fib-4 (<1.3 vs ≥1.3) | **17.76 (1.77)** | **3.25 (0.89)** | **11.35 (1.74)** | 0.78 (0.41) | **0.011** | **0.009** | **10.62 (0.20)** | **0.68 (0.07)** | **5.19 (0.22)** | **-0.20 (0.06)** | **<0.001** | **<0.001** |
| GFR (≥90 vs ≤90) per unit decrease | **0.49 (0.06)** | **0.19 (0.04)** | **0.12 (0.06)** | **0.17 (0.03)** | **<0.001** | 0.73 | **0.32 (0.01)** | **0.14 (0.00)** | **0.15 (0.01)** | **0.06 (0.00)** | **<0.001** | **<0.001** |

## Discussion

In two large independent cohorts of ambulatory adults, we found that women compared to men consistently demonstrated a larger magnitude and an earlier onset of vascular sensitivity in the setting of metabolic disease stress of varying types – including both systemic as well as organ-specific sources of metabolic stress. Sex differences in the slope of SBP increase were especially pronounced in relation to increasing levels of metabolic stress within the normal range – suggesting a greater sensitivity in women during the transition from metabolic health to metabolic disease. In our large-sized ambulatory patient cohort, sex differences in slope of SBP elevation persisted across both normal and abnormal metabolic stress measurement values suggesting that women retain a greater vascular sensitivity to systemic and organ-specific stress from the earliest through the latest stages of metabolic disease. Extending from previous reports demonstrating excess CVD risk in women compared to men with overt metabolic disease, our findings suggest that efforts to mitigate vascular disease risk in women should include targeting metabolic stressors – and that such targeting should begin early on, well prior to the onset of clinically evident metabolic disease.

Our study limitations included the inability to examine more detailed measures of vascular health or metabolic stress beyond the measures that were readily available across both our community and our ambulatory patient cohorts. The cross-sectional study design also precludes ability to determine temporality of associations. Notably, regarding our observed associations of metabolic stress measures with SBP, bidirectional or reverse causation is biologically plausible albeit much more so for some measures (e.g. eGFR) than for others (e.g. HbA1c, FIB-4). Future prospective and longitudinal studies are needed to clarify temporality and directionality of our observed associations.

In conclusion, we found evidence of vascular sensitivity in the setting of even the mildest forms of pre-clinical metabolic stress; notably, this vascular sensitivity was not specific to any given systemic or organ-derived metabolic stress. Moreover, the finding was especially pronounced in women compared to men and so may represent an important precursor to sex differences in cardiovascular outcomes. Notwithstanding the need for further work to examine underlying mechanisms, these results underscore the importance of ongoing endeavors to involve metabolic interventions as part of efforts to mitigate vascular risks in both women and men.

## Data Availability

The NHANES data were acquired from and are available to the scientific community through https://www.cdc.gov/nchs/nhanes/index.htm. Data was accessed on 4/1/2024 through R Package nhanesA. The internal data that support the findings of this study are available on request from the corresponding author.

