## Supplemental Table 1 for "Multi-Organ System Metabolic Stress and Sex-Divergent Vascular Associations"

| **Condition** | **ICD 10 Diagnosis Codes** |
| --- | --- |
| **Hypertension** | I10, I11*, I12*, I13*, I15*, I16*, I67.4, I97.3 |
| **Hyperlipidemia** | E75.6, E78* |
| **Diabetes** | E10*-E14* |
| **Coronary Artery Disease** | I21*, I22*, I25.2 |
